# The Effect of Legalizing Online Sports Gambling on Population Mental Health

**DOI:** 10.64898/2026.05.06.26352568

**Authors:** Nolan M. Kavanagh, Jacob C. Jameson, Harold A. Pollack, Nathaniel J. Glasser

## Abstract

**Importance:** The rapid rise of online sports gambling in the U.S. has been associated with financial harms, raising concern that it may adversely affect population mental health.

**Objective:** To estimate the causal effect of state legalization of online sports gambling on population mental health, including a range of self-reported and registry-based outcomes.

**Design, Setting, and Participants:** Repeated cross-sectional study using nationally representative Behavioral Risk Factor Surveillance System (BRFSS) data from 2014–2025 and registry-based mortality records from 2012–2024. We leveraged state-level variation in the legalization of online sports gambling and applied a stacked difference-in-differences with event study design. The analytic sample included 4,660,948 BRFSS respondents and mortality records for virtually all state-years. We estimated effects on all adults and several higher-risk subgroups, including men, young men, and men with lower educational attainment.

**Exposure:** State legalization of online sports gambling.

**Main Outcomes and Measures:** Self-reported outcomes included poor mental health days, depressive disorder diagnoses, ever binge drinking, number of binge drinking episodes, and marijuana use. Registry-based outcomes included suicide mortality and alcohol-induced mortality per 100,000.

**Results:** Among 4,660,948 BRFSS respondents, 48.7% were men, 40.2% had no more than a high school education, and the mean age was 47.6 years. Legalization of online sports gambling had no discernible effect on poor mental health days of all U.S. adults (–0.01 days; 95% CI, –0.16 to 0.14; P=0.88), depressive disorder diagnoses (0.1 percentage points; 95% CI, –0.7 to 0.9; P=0.84), binge drinking, binge drinking episodes, or marijuana use. Meanwhile, mean suicide mortality was 14.1 per 100,000 and mean alcohol-induced mortality was 12.2 per 100,000. Legalization did not affect adult suicides (0.13 deaths per 100,000; 95% CI, –0.71 to 0.97; P=0.76) or alcohol-induced mortality (1.08 deaths per 100,000; 95% CI, –0.58 to 2.73; P=0.21). Results were null among men and higher-risk subgroups of men.

**Conclusions and Relevance:** The legalization of online sports gambling has not produce detectable population-level changes in a range of mental health outcomes, including reported symptoms, diagnoses, substance use, and registry-based mortality due to suicide or alcohol, in up to 3 years of follow-up. These findings suggest that although online sports gambling may cause financial harm and severe distress for some individuals, legalization has not produced measurable average changes in population mental health over the observed follow-up period.

**Key points:** *Question:* Has the legalization of online sports gambling affected population-level mental health, including symptoms, diagnoses, substance use, suicides, and alcohol-induced mortality?

*Findings:* In this repeated cross-sectional study that applied a difference-in-differences design to more than 4.6 million individual-level survey responses and mortality records, the legalization of online sports gambling from 2018–2024 did not affect reported poor mental health days, depressive disorders, binge drinking, marijuana use, suicide mortality, or alcohol-induced mortality. Results were similar among men and higher-risk subgroups of men.

*Meaning:* The legalization of online sports gambling has not produced detectable population-level changes in a broad range of mental health outcomes in up to 3 years of follow-up.

## Introduction

In 2018, the U.S. Supreme Court struck down a federal ban on sports gambling.^1^ Since then, 38 states and the District of Columbia (D.C.) have legalized sports gambling, with 31 permitting it online. Participation has grown dramatically over time. In 2024, Americans placed nearly $150 billion in legal sports bets,^2^ and 22% of U.S. adults — including 48% of men ages 18 to 49 — reported having used a sports betting app.^3^

The rollout of sports betting has been shown to worsen population-level financial well-being, with decreased long-term investments, lower credit scores, and higher rates of catastrophic outcomes such as bankruptcies and collections.^4,5^ These effects are especially pronounced among already financially constrained households.^4^ Roughly 20% of betters in 2024 reported struggling to meet financial obligations because of their losses.^3^

Although betting may provide entertainment, the apparent financial consequences raise the prospect that sports betting may threaten the mental health of populations.^6^ These effects might proceed through the direct stress of financial losses on betters who struggle to meet financial obligations, or via downstream effects, if depleted assets preclude investments in betters’ health.^7^ To that end, sports gambling has also been associated with worse mental health at the individual level,^7–12^ albeit not in all studies.^13^ Sports betting may also have adverse consequences for spouses or other household members affected by betters’ behaviors.^14–16^ However, these studies are typically cross-sectional and lack well-identified counterfactuals necessary to establish causal relationships.

Based on existing evidence, then, it is less clear whether the recent rise in sports gambling has caused observable population-level effects on mental health, substance use, and related outcomes. Recent descriptive work suggests a rise in concerns about gambling addiction,^1^ and troublingly, sports betting has been tied to increased incidence of intimate partner violence.^14,15^ Among studies that have used rigorous designs to assess the mental health consequences of sports gambling, the effects were either uneven across outcomes and demographic groups, limited to the early years after legalization, or greater in apparent magnitude than the number of households that have engaged in sports betting.^17–19^

We leverage state-level variation in the legalization of online sports gambling and apply a rigorous stacked difference-in-differences design to test the causal effect of these policies on population mental health. In doing so, we use millions of person-years of data and multiple data sources to interrogate a range of mental health outcomes, including symptoms, diagnoses, substance use, and mortality. We also examine the effects on several subgroups at higher risk of gambling-related distress. The findings have important implications for policy makers looking to protect the well-being of populations.

## Methods

### Data and Outcomes

Our data came from (1) the pooled Behavioral Risk Factor Surveillance System (BRFSS), a large repeated cross-sectional study of the health of U.S. adults, from 2014 to early 2025, for self-reported symptoms, diagnoses, and substance use; and (2) the Center for Disease Control and Prevention’s (CDC) Wide-ranging ONline Data for Epidemiologic Research (WONDER), a registry of U.S. mortality by underlying cause from 2014 to 2024. The two complement each other by providing outcomes that capture diverse facets of mental health, one relying on self-report, the other on death records from state registries.

BRFSS outcomes were all self-reported and included (1) the number of days in the past 30 days when a respondent’s mental health was not good due to stress, depression, or problems with emotions; (2) whether respondents were ever told they have a depressive disorder (coded as yes or no); (3) ever binge drinking in the past 30 days (defined as 5 or more drinks for men or 4 or more drinks for women on a single occasion; coded as yes or no); (4) number of binge drinking episodes in the past 30 days (5) and use of marijuana or cannabis in the past 30 days (available in select states and years starting in 2016 [**eFigure 1**]; coded as yes or no). Together, these outcomes broadly include symptoms that may directly reflect the distress that could result from gambling, as well as diagnoses and substance use that may reflect health care-seeking and coping strategies.

In the BRFSS analyses, respondents were excluded listwise from all analyses if missing information on age (0%), sex (<1%), or education (<1%). Those missing data on a given outcome were omitted pairwise from those analyses; missingness was 2% for days of poor mental health, 1% for depressive disorders, 8% for binge drinking, and 14% for marijuana in the states and waves when the marijuana module was included. All BRFSS estimates were weighted to be nationally representative using survey weights.

Our CDC registry outcomes were (1) unadjusted deaths by suicide per 100,000 people in each state, and (2) all alcohol-induced deaths, including poisonings, per 100,000 people. For mortality analyses, we had nearly complete data by state and year, except in rare states-years for small subgroups when there were fewer than 10 deaths in a state-year. All estimates were weighted by state population to be representative of the U.S.

### Exposure

Our exposure is a state’s legalization of online sports gambling. Since 2018, 29 states plus D.C. have done so (**Figure 1**). We focused on online betting since it has had a greater impact on financial well-being than retail betting.^4,5^ Nevada was excluded from all analyses since it legalized gambling before 2018 and is “always treated.” The 8 states with retail-only sports gambling were used as control states up until the date when they allowed retail gambling, after which they were excluded. Legalization dates were taken from Baker et al, who observed deposits to online gambling platforms in each state, allowing them to confirm that legalization was actually accompanied by observable gambling activity.^4^

**Figure 1.**
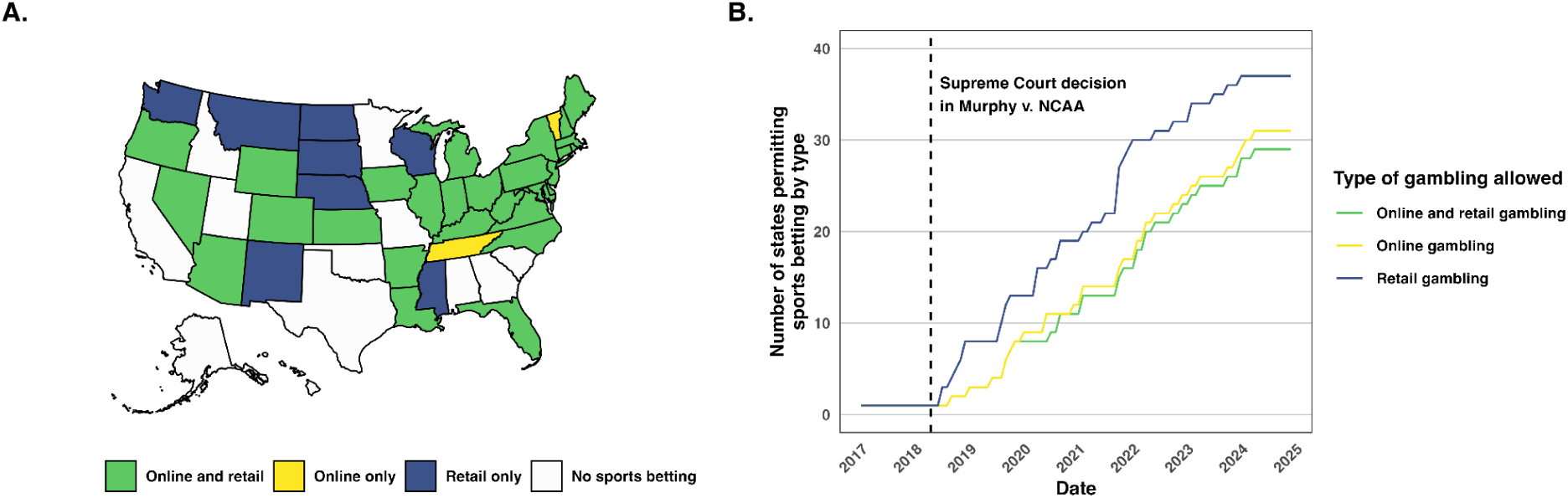
Trends in State Legalization of Online Sports Gambling. **Notes: Panel A** shows the states that have legalized sports gambling by type as of early 2025, the last period for which we have data. **Panel B** shows the trends in the number of states that have legalized online, retail, or both online and retail sports betting over time.

### Study Design

We used a stacked difference-in-differences with event study design to test the causal effect of state legalization of online sports gambling on population mental health (**eMethods**). This approach is akin to a standard difference-in-differences except that it combines multiple sets of treated states that passed a policy of interest in different years. To construct it, we take the cohort of states that legalized online sports gambling in a given quarter and construct a set of control states that never do so or will not have done so during the given post-period. Then, we “stack” all legalization cohorts into a single model. This approach prevents inappropriate comparisons between newly treated and already-treated states, which can introduce bias into pooled difference-in-differences models.^20,21^

For the BRFSS outcomes, we used a pre-period of 4 years and post-period of 2.5 years (except for marijuana use, which used a pre-period to 2 years since marijuana use was first assessed by the BRFSS in 2016). BRFSS outcomes are summarized by quarter. For the CDC outcomes, we used a pre-period of 6 years and post-period of 3 years. These outcomes are summarized by year. The pre- and post-period lengths were chosen to balance the competing desires to assess for longer-term effects while minimizing the number of states with recent legalizations that would not have full post-periods.

Effects were estimated using OLS models with fixed effects for state-by-stack to account for state policy environments and quarter-by-stack to account for national trends. The interaction of stack with the unit and time in the fixed effects is essential in this design since units can be included in multiple “stacks.” Models using the BRFSS data also included fixed effects for respondent birth year to account for generational differences in mental health. Standard errors were clustered by state since policy decisions to legalize sports gambling were made at that level. We estimated two versions of each model, one on a balanced panel of 22 treated units (with the full number of post-period years) and another on an imbalanced panel of 30 (where 8 states have less than full follow-up).

We start by estimating the population-wide effects of legalized online sports gambling on all U.S. adults. Given that men, especially younger ones and those in more economically precarious households, are more financially affected by sports gambling,^4,5^ we also estimated the effects on several subgroups at higher risk of distress: in the BRFSS, men, men born 1973–1997 (who would have been ages 21–45 in 2018 when gambling was initially legalized),^1^ and men with no more than a high school education. In mortality data, we estimated subgroups of men and men under 45 (since the data does not include detailed demographic information, including birth year or educational attainment).

### Additional Analyses

We conducted several additional analyses to assess the robustness of our findings and situate them in the existing literature. Since not all states fielded the BRFSS marijuana module in all years (**eFigure 1**), our estimates could be biased by selection in the included states, especially the event studies. To reassure against any bias, we re-estimated the main model for all adults using states with more complete marijuana data, i.e. at least 5 waves.

Dasgupta and Ghimire identified large effects on episodes of binge drinking conditional on binge drinking at least once among men under 35,^19^ so we replicated this analysis using the same outcome and age group. Models were otherwise as described above. And Couture et al identified heterogeneous effects, with men ages 18–24 experiencing a lower probability of having at least one bad mental health day, and men ages 30–34 experiencing a higher probability.^17^ We replicated this analysis using the same outcome and age groups.

All analyses were performed in R (v. 4.3.1) in the “lfe” package (v. 3.1.1). This study did not require institutional review board approval since it used public, de-identified data. All replication materials are publicly available at the Harvard Dataverse (link TBD).

## Results

### Self-Reported Mental Health Outcomes

From 2014 to early 2025, 4,765,902 respondents were included in the final BRFSS sample. A weighted 48.7% were men, 40.6% had no more than a high school education, and the mean (SD) age was 47.7 (18.1) years. Among all adults, the mean (SD) number of bad days of mental health in the past 30 days was 4.3 (8.3), and 19.0% reported having a depressive disorder. Meanwhile, 16.2% reported binge drinking at least once during the past month, with a mean (SD) of 0.7 (3.1) episodes of binge drinking (including non-binge drinkers), and 12.3% reported ever using marijuana in the past month. The trends over time, as well as the rates for several subgroups, are presented in **Figure 1**.

Among all adults, state legalization of online sports gambling had no apparent effect on the mean number of reported bad mental health days (–0.01 days; 95% CI, –0.16 to 0.14, P=0.88), rates of depressive disorders (0.1 percentage points; 95% CI, –0.7 to 0.9, P=0.84), rates of binge drinking in the past month (0.1 percentage points; 95% CI, –0.7 to 0.9, P=0.84), episodes of binge drinking in the past month (0.1 percentage points; 95% CI, –0.7 to 0.9, P=0.84), and rates of using marijuana in the past month (0.1 percentage points; 95% CI, –0.7 to 0.9, P=0.84). The pooled estimates are shown in **Figure 2**.

**Figure 2.**
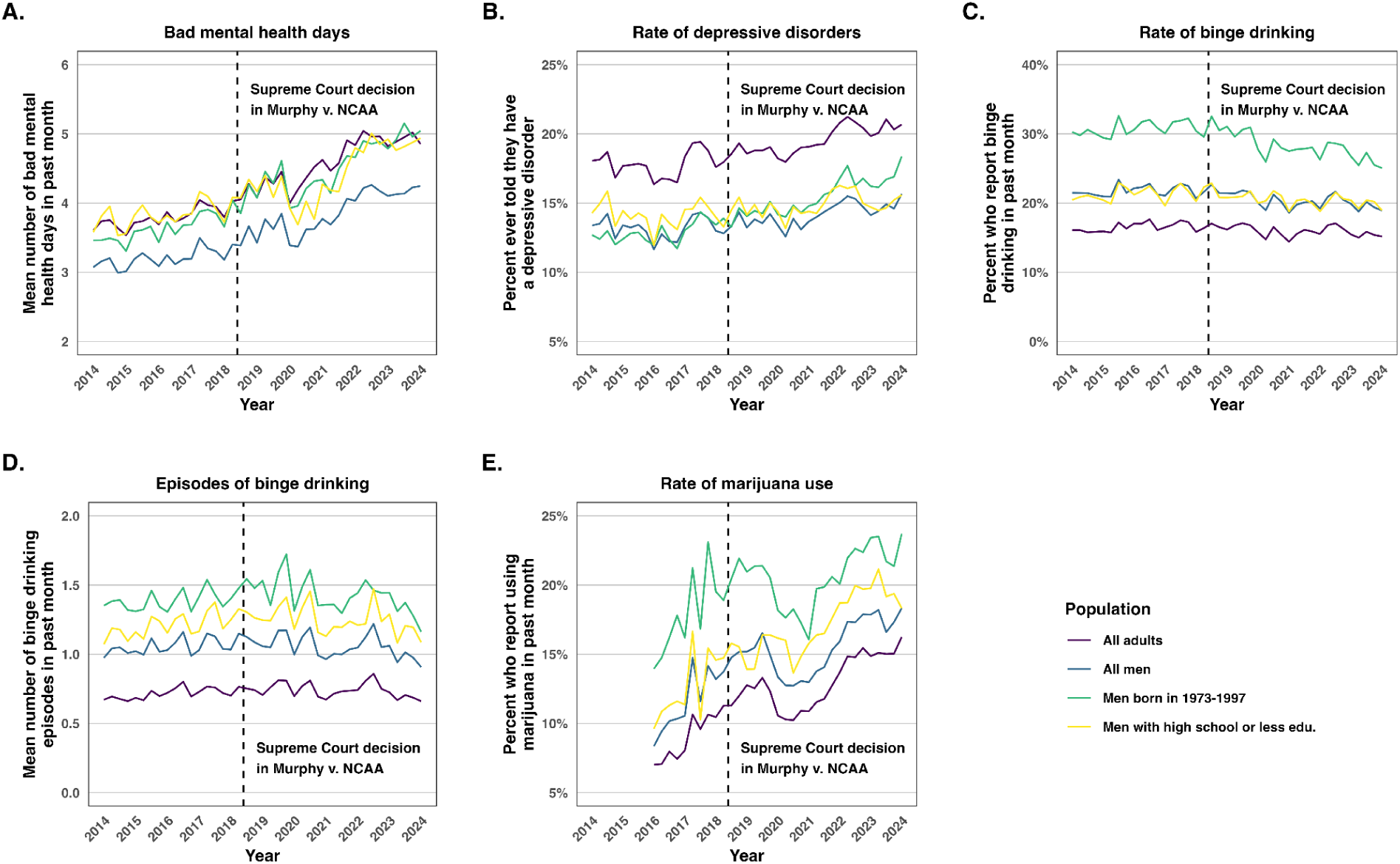
Trends in Mental Health and Substance Use Among U.S. Adults, 2014–2024. **Notes:** Quarterly estimates from the Behavioral Risk Factor Surveillance System for all U.S. adults and indicated subgroups, weighted to be nationally representative. **Panel A** shows the mean number of days in the past 30 days when respondents reported that their mental health was not good. **Panel B** shows the percentage of respondents who reported ever being told they have a depressive disorder (i.e. a lifetime diagnosis). **Panel C** shows the percentage of respondents who reported binge drinking in the past 30 days (defined as 5 or more drinks for men or 4 or more drinks for women on a single occasion). **Panel D** shows the mean number of binge drinking episodes in the past 30 days. **Panel E** shows the percentage of respondents who reported using marijuana or cannabis in the past 30 days (available select states and years after 2016, as indicated in **eFigure 1**). The vertical dashed line marks the 2018 U.S. Supreme Court decision in *Murphy v. NCAA*, which struck down the federal ban on sports gambling. We include data for early 2025 in our models but do not show them here because the quarter is incomplete. **Abbreviations:** NCAA, National College Athletic Association.

The event studies are presented in **Figure 3**. They rule out even small effects on each outcome up to 2.5 years after the legalization of online sports betting. They also reassure against non-parallel pre-trends between treated and control states that would threaten causal inference. One exception is marijuana use, which has an apparent violation of parallel pre-trends, suggesting that our control states are an imperfect counterfactual for treated states. As a result, we caution against a strong causal claim for this outcome.

**Figure 3.**
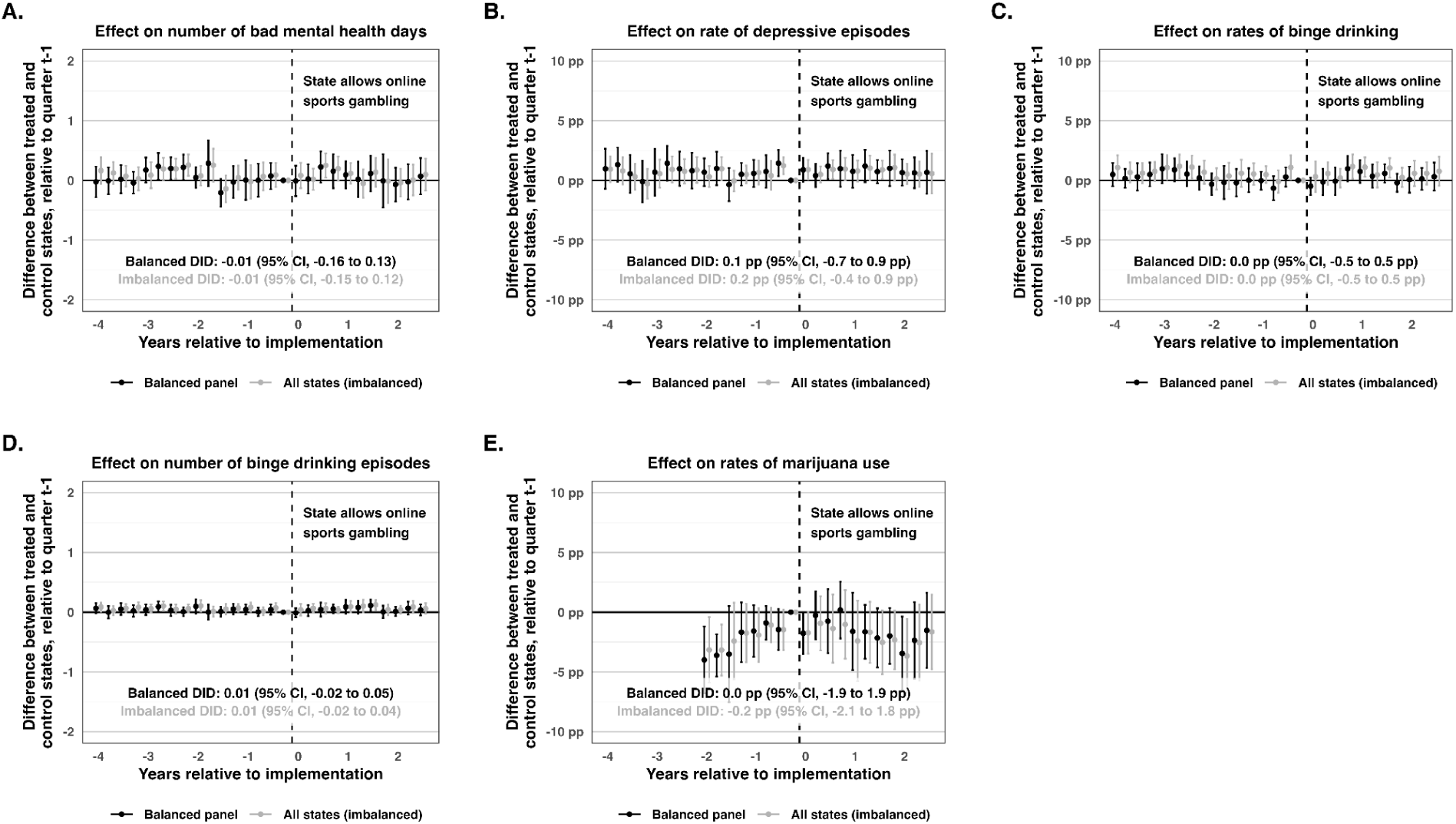
Event Studies for the Effect of Legalizing Online Sports Gambling on Population Mental Health Among All U.S. Adults. **Notes:** Based on data from the Behavioral Risk Factor Surveillance System from 2014–2025. Each panel presents a stacked difference-in-differences (DID) event study comparing outcomes between treated and control states, relative to the quarter before legalization of online sports gambling. We estimated two models for each outcome: a balanced panel of 22 treated states with a full 2.5 years of post-legalization data (excluding Nevada, which legalized online gambling before 2018) and an imbalanced panel of all 30 treated states (with some having incomplete post-legalization data). Estimates for the pooled difference-in-differences model are provided. **Panel A** examines the mean number of days of poor mental health in the past 30 days. **Panel B** examines rates of respondents ever told they have a depressive disorder. **Panel C** examines rates of ever binge drinking in the past 30 days. **Panel D** examines the number of binge drinking episodes in the past 30 days. **Panel E** examines rates of marijuana use in the past 30 days. All models included fixed effects for state-by-stack, quarter-by-stack, and respondent birth year. 95% confidence intervals clustered at the state level are provided. Estimates used survey weights. **Abbreviations:** NCAA, National Collegiate Athletic Association; pp, percentage point; DID, difference-in-differences.

**Figure 4.**
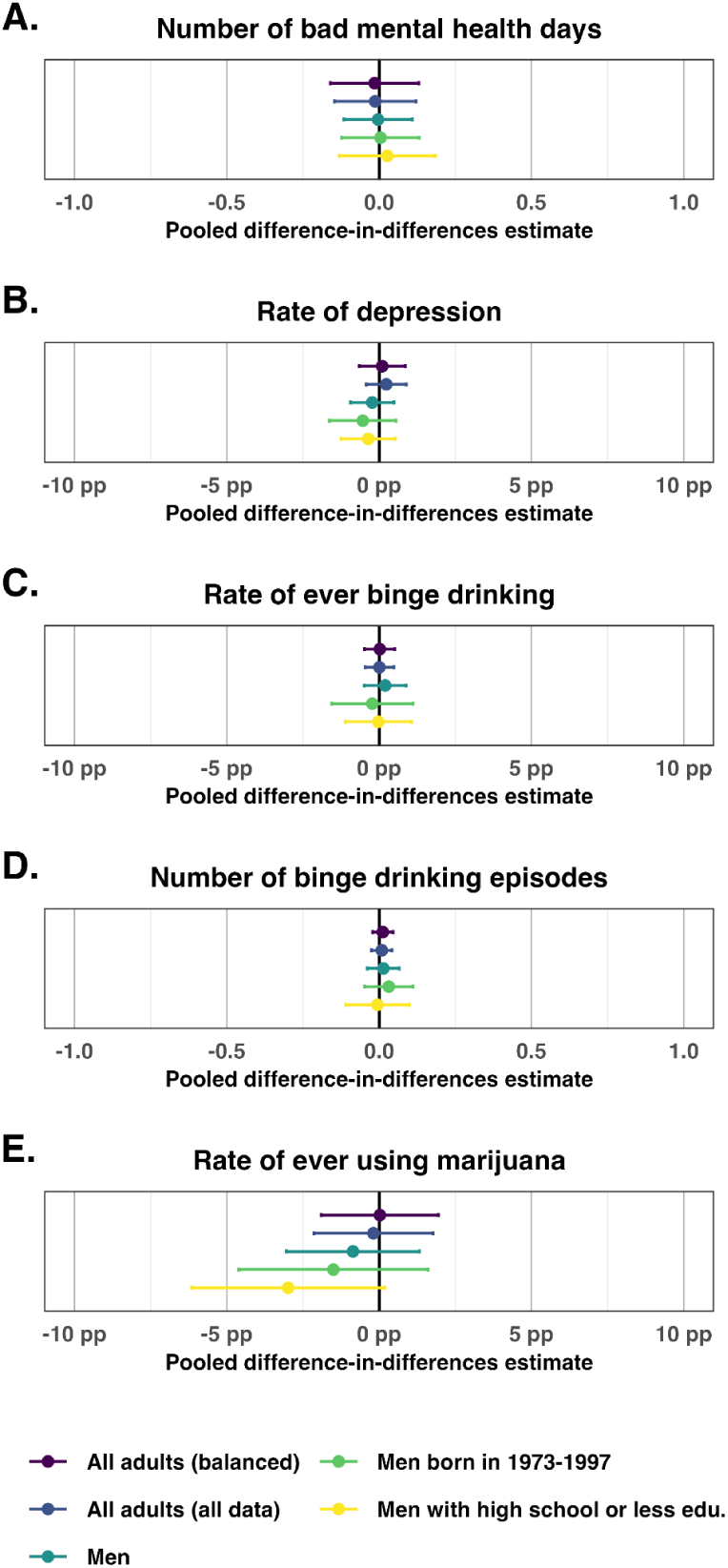
Pooled Difference-in-Differences Estimates for the Effect of Legalizing Online Sports Gambling on Population Mental Health. **Notes**: Based on the Behavioral Risk Factor Surveillance System from 2014 to early 2025. Each panel presents the estimate from a pooled, stacked difference-in-differences model for the indicated outcome and subgroup. For all adults, we present estimates using both a balanced and imbalanced panel of treated states; all subgroup analyses required balanced panels. 95% confidence intervals clustered at the state level are provided. **Panel A** is measured in days; **Panels B, C,** and **E** are measured in percentage points; and **Panel D** is measured in episodes. All models included fixed effects for state-by-stack, quarter-by-stack, and respondent birth year, and all estimates used survey weights. **Abbreviations:** pp, percentage points; edu., education.

In subgroup analyses, there were no effects on an imbalanced panel of all adults (that is, a panel that includes states that do not have 2.5 years of post-legalization data), men, men born from 1973–1997, and men with no more than a high school education for any of the five self-reported mental health outcomes in the BRFSS (**Figure 3**).

In additional analyses, restricting the marijuana analysis of all adults to states with more complete marijuana data produced similar estimates (**eFigure 2**). In our replication of Dasgupta and Ghimire, we did not find any clear effects on the number of binge drinking episodes, conditional on having at least one, among men under 35 (**eFigure 3**). And in our replication of Couture et al, we did not find any effects on the probability of having at least one bad mental health day among men ages 18–24 or men ages 30–34 (**eFigure 4**).

### Registry-Based Mortality Outcomes

From 2012–2024, there were 598,539 suicides and 516,878 alcohol-induced deaths in CDC WONDER mortality records. The mean suicide rate was 14.1 deaths per 100,000 population among all adults, 22.4 among men, and 17.4 among men younger than 45 years (**Figure 5**). The mean alcohol-induced mortality rate was 12.2 deaths per 100,000 population among all adults, 17.2 among men, and 5.0 among men under 45.

**Figure 5.**
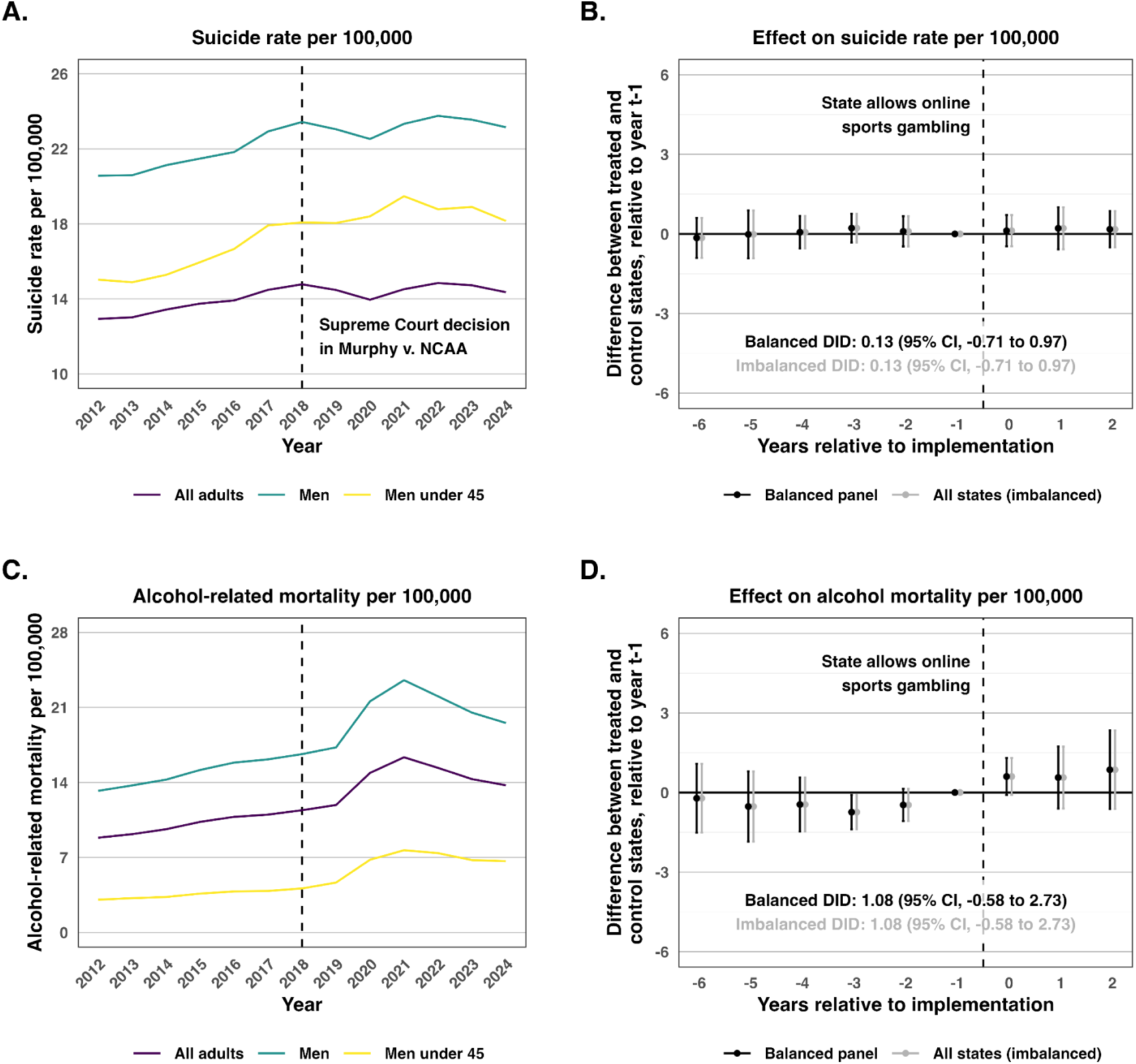
Trends and Event Studies for the Effect of Legalizing Online Sports Gambling on Suicide and Alcohol-Induced Mortality, 2012–2024. **Notes**: Based on CDC death records from 2012 to 2024. **Panels A** and **C** present trends in unadjusted rates of suicides and alcohol-induced deaths per 100,000 population, respectively, for all U.S. adults, men, and men younger than 45 years. **Panels B** and **D** present stacked event studies for all U.S. adults, comparing the differences between treated and control states for each year, relative to the year before online sports gambling was legalized. Estimates for the pooled difference- in-differences model are provided. Models included fixed effects for state-by-stack and year-by-stack. Estimates were weighted by the population in each state-year. 95% CIs clustered at the state level are provided. **Abbreviations:** DID, difference-in-differences.

In stacked difference-in-differences models, legalization of online sports gambling had no effect on suicide mortality among all adults (0.13 deaths per 100,000; 95% CI, –0.71 to 0.97; P=0.76) (**Figures 5A–5B**). Event studies showed no evidence of pre-trend violations or dynamic effects up to 3 years after legalization. Similarly, there was no observable effect on alcohol-induced mortality in pooled models (1.08 deaths per 100,000; 95% CI, –0.58 to 2.73; P=0.21) (**Figures 5C–5D**). The event studies may suggest some increasing alcohol-induced mortality, although the coefficients remain null in each year and the points have the same trajectory as the two pre-legalization years. The results were similar in analyses on imbalanced panels of states with all available follow-up.

In subgroup analyses, legalization had no effect on suicide mortality among men (0.32; 95% CI, –1.24 to 1.88) or men younger than 45 years (0.15; 95% CI, –1.26 to 1.56) (**eFigure 5**). Estimates for alcohol-induced mortality were similarly null among men (1.69; 95% CI, –0.54 to 3.93) and men younger than 45 years (0.78; 95% CI, –0.55 to 2.10) (**eFigure 6**). Akin to the models among all adults, the event studies suggested worsening trends in alcohol-related mortality but were confounded by potential pre-trend violations.

Overall, registry-based mortality analyses did not indicate clear population-level mental health harms following the legalization of online sports gambling.

## Discussion

Despite financial harms and growing public concern,^4–6^ the legalization of online sports betting has not had measurable population-level effects on the mental health of U.S. adults in this nationally representative study using a rigorous causal design. Across several outcomes spanning reported symptoms, clinical diagnoses, substance use, and mortality, we estimated null effects among all adults and several subgroups at highest risk of gambling-related distress, including men, young men, and men with lower educational attainment. Event studies showed no dynamic effects up to 3 years after legalization.

How should we interpret these findings? As with any null result, we cannot rule out very small effects. However, the BRFSS analyses allow us to rule out meaningfully large ones. Among all adults, the upper bound of the 95% CI for bad mental health days excludes a population-wide effect of 0.14 or more days per month. If the mental health burden is primarily shouldered by betters, who comprise 22% of U.S. adults,^3^ then we can rule out an average effect on bettors larger than 0.6 additional days of poor mental health per month.

Among men ages 21–45, whose betting participation approaches 48%,^3^ the bound is even tighter at roughly 0.3 days per bettor. These bounds do not exclude the possibility that a small number of individuals experience severe gambling-related distress. However, any such effects were neither large enough nor prevalent enough to shift the population means, even among higher-risk subgroups. The mortality outcomes were also null but less precise. The event studies for alcohol-related mortality, in particular, suggested worsening trends, although we lack the statistical power for firm conclusions in either direction.

It is worth considering potential explanations for our null findings. We suspect that a major reason is simply that, while many Americans engage in sports betting, few do so to a degree that would likely cause mental distress. For example, Baker et estimated that 8% of households ever placed an online sports bet from 2010–2023; among them, the average deposits were about $400 per year, or 0.7% of their income. Among the highest tercile of betters, which would correspond to roughly 2% of households, deposits averaged roughly $1,200, or 1.7% of income. This is akin to an effective 1–2% pay cut for the most active betters. To be sure, participation in betting has increased since 2023, but put into context, studies on the minimum wage using high-quality causal designs have often struggled to identify mental health impacts of much larger relative changes in income.^22–26^ The number of households with even more debilitating betting is a yet smaller fraction and unlikely to shift population means to a sufficient degree that could be observed in our design.

Our results diverge from several recent studies on sports gambling and mental health using the same datasets. Dasgupta and Ghimire reported a 10% increase in binge drinking episodes among men under 35 years due to online sports gambling.^19^ By contrast, we estimated precise nulls for binge drinking in all populations we examined, including a direct replication with their preferred outcome. Also of note, Dasgupta and Ghimire did not find effects on the 6 other measures of alcohol consumption they examined.^19^ Couture et al reported heterogeneous effects by age, with men ages 18–24 experiencing an 11% decrease in the likelihood of poor mental health after legalization, while men ages 30–34 experiencing a 10% increase, and no effects among women.^17^ Of note, their study was limited to 8 states that legalized before the COVID-19 pandemic with a short follow-up period. Our replication did not show any evidence of effects in either group of men. Lastly, Orujov et al reported effects on suicide mortality that we could not replicate in our analyses with CDC WONDER data.^18^ It is not immediately obvious why our results differ. One possibility is that many of these studies are based on the early states that legalized online sports gambling and used shorter post-legalization follow-up. It may be that as more states have implemented the policy for more time, those early signals have not persisted.

This study has limitations. First, our outcomes do not capture all dimensions of mental health that gambling might impact. For example, we do not observe gambling-specific distress, relationship strain, or subclinical anxiety that falls below the threshold of a “bad mental health day.” Second, the BRFSS outcomes rely on self-report, and the populations most affected by online sports betting — young adult men — may be more likely to downplay mental distress or problem behaviors and less likely to participate in health surveys.^27–29^ That said, we also find null results for the registry-based outcomes, albeit less precisely estimated. Third, the post-legalization follow-up period of over 2 years, while longer than prior studies, may still be insufficient if the mental health consequences of gambling accumulate slowly. Finally, our design estimates the reduced form — or population-wide effect — of legalized online sports gambling as a policy; it does not identify the effect of gambling on specifically those who participate, which may differ.

Despite these limitations, the breadth and consistency of the null findings across multiple outcomes, data sources, and subgroups are informative. While problem gambling and poor mental health have been associated at the individual level in cross-section,^8–10^ the legalization of online sports gambling has (yet) not produced a detectable shift in the mental health of populations. This is not to say that gambling is without harm. Sports gambling has had demonstrable financial consequences,^4,5^ and there is suggestive evidence of increased intimate partner violence.^14,15^ There may be other normative or empirical reasons to restrict online gambling. At the same time, amidst rising loneliness, mental distress, and suicidality among young men,^30–32^ it appears less likely that online sports gambling is a major driver of these trends. Attributing them to sports gambling may mean that policy makers miss the opportunity to identify and address drivers more likely to have a durable impact.

## Data Availability

Data are publicly available. All replication materials will be publicly posted in the Harvard Dataverse upon publication of the paper.

---

**eTable 1:** Sport Betting Legalization Dates.

| State | Online Launch Date | Retail Launch Date |
| --- | --- | --- |
| Arizona | 2021-09-09 | 2021-09-09 |
| Arkansas | 2022-03-06 | 2019-07-01 |
| Colorado | 2020-05-01 | 2020-06-17 |
| Connecticut | 2021-10-19 | 2021-09-30 |
| Delaware | 2023-12-27 | 2018-06-05 |
| Florida | 2023-11-07 | 2023-12-07 |
| Illinois | 2022-03-05 | 2020-03-09 |
| Indiana | 2019-10-03 | 2019-09-01 |
| Iowa | 2019-08-15 | 2019-08-15 |
| Kansas | 2022-09-01 | 2022-09-01 |
| Kentucky | 2023-09-28 | 2023-09-07 |
| Louisiana | 2022-01-28 | 2021-10-06 |
| Maine | 2022-05-02 | 2022-05-02 |
| Maryland | 2022-11-23 | 2021-12-09 |
| Massachusetts | 2023-03-10 | 2023-01-31 |
| Michigan | 2021-01-22 | 2020-03-11 |
| Mississippi | – | 2018-08-01 |
| Montana | – | 2020-03-11 |
| Nebraska | – | 2023-06-22 |
| Nevada | 2010-01-01 | 1949-01-01 |
| New Hampshire | 2019-12-30 | 2020-08-12 |
| New Jersey | 2018-08-01 | 2018-06-14 |
| New Mexico | – | 2018-10-16 |
| New York | 2022-01-08 | 2019-07-16 |
| North Carolina | 2024-03-11 | 2021-03-18 |
| Ohio | 2023-01-01 | 2023-01-01 |
| Oregon | 2019-08-27 | 2019-08-27 |
| Pennsylvania | 2019-05-01 | 2018-11-15 |
| Rhode Island | 2019-09-14 | 2018-11-26 |
| South Dakota | – | 2021-09-09 |
| Tennessee | 2020-11-01 | – |
| Vermont | 2024-01-11 | – |
| Virginia | 2021-01-21 | 2021-01-21 |
| Washington | – | 2021-09-09 |
| Washington, D.C. | 2020-05-28 | 2020-08-31 |
| West Virginia | 2018-12-01 | 2018-09-01 |
| Wisconsin | – | 2021-11-30 |
| Wyoming | 2021-09-01 | 2021-09-01 |

**Notes:** – indicates that the betting type is not available in that jurisdiction. Nevada is excluded from all models since its legalization of both forms of gambling predates 2018, i.e. it is “always treated.” Mississippi, Montana, and Washington allow online betting only when on the premises of a retail location; as such, they are treated as retail betting states in our analyses. All dates were taken from Baker et al’s working paper from 2024, who were able to confirm the legalization dates by observing deposits into sports betting apps.

**eFigure 1.**
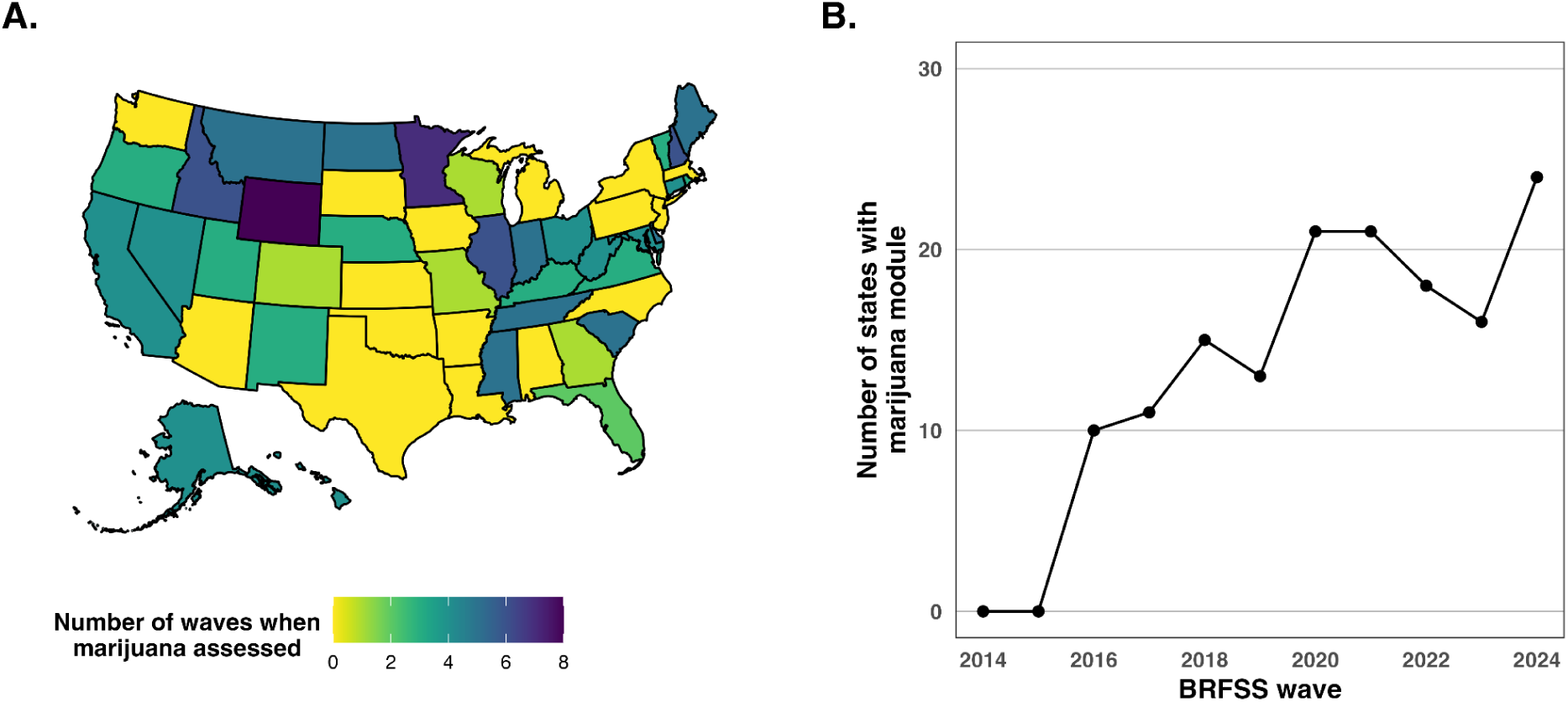
Inclusion of Marijuana Module in the BRFSS Over Time. **Notes:** The BRFSS began assessing marijuana use in 2016 as an optional module. **Panel A** depicts the number of years that each state included the marijuana module, and **Panel B** depicts the number of states that included it by survey wave. Because of this missingness, estimates from the marijuana models should be interpreted with some caution.

**eFigure 2.**
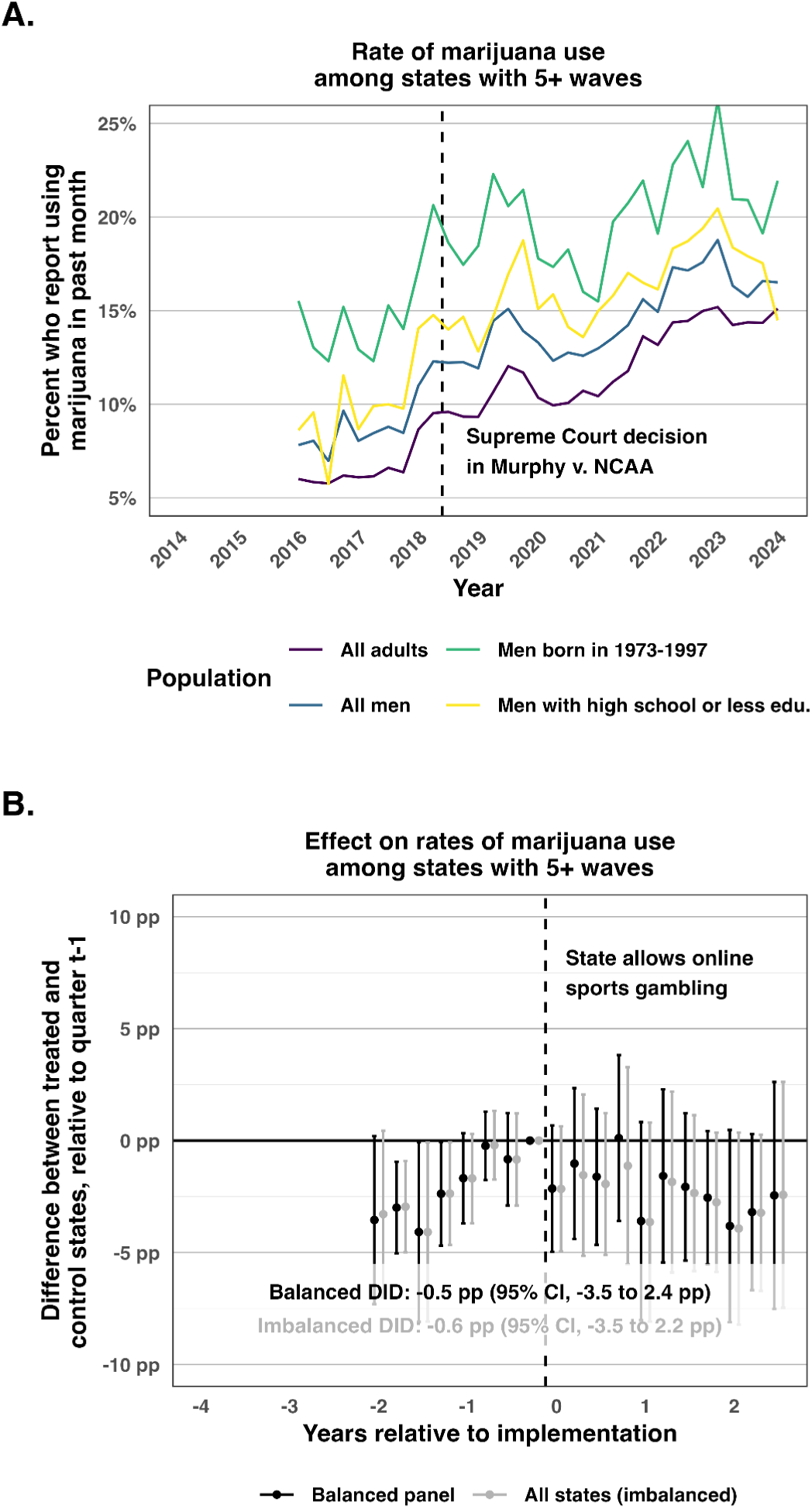
Marijuana Analyses on States with 5+ Waves. **Notes:** Robustness analyses with a re-estimation of Figures 2E and 3E restricted to states with more consistent inclusion of the marijuana BRFSS module, i.e. states with at least 5 waves (of a possible 8). The results are similar to the main models, reassuring against bias due to selection in which states fielded the marijuana module in which years.

**eFigure 3.**
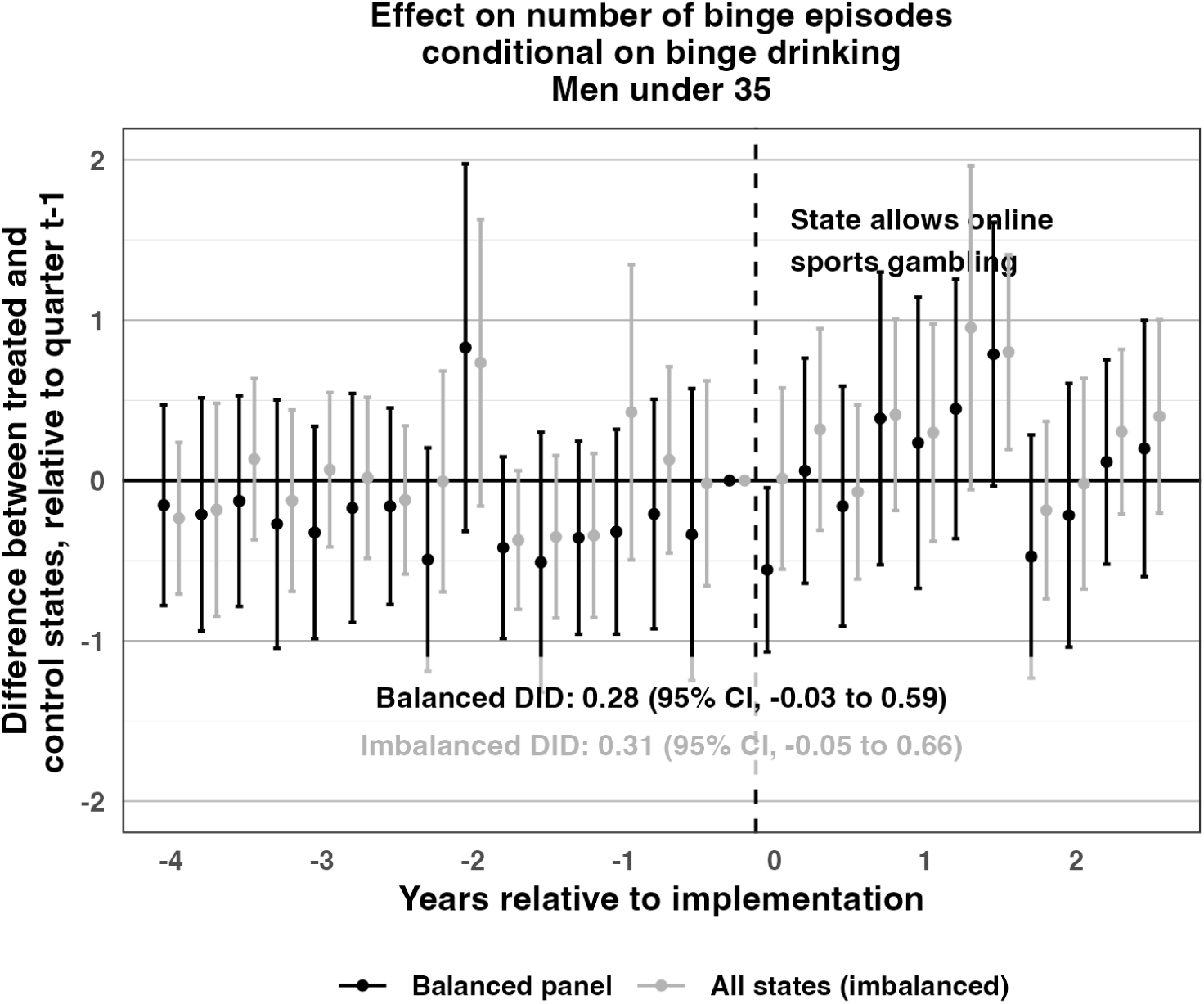
Replication of Dasgupta and Ghimire (2026). **Notes:** We replicated the preferred outcome and age group of Dasgupta and Ghimire (2026), i.e. number of binge drinking episodes conditional on having at least one among men under 35. Models otherwise have the same specifications as our main models. We did not find clear evidence that online sports gambling increased binge drinking.

**eFigure 4.**
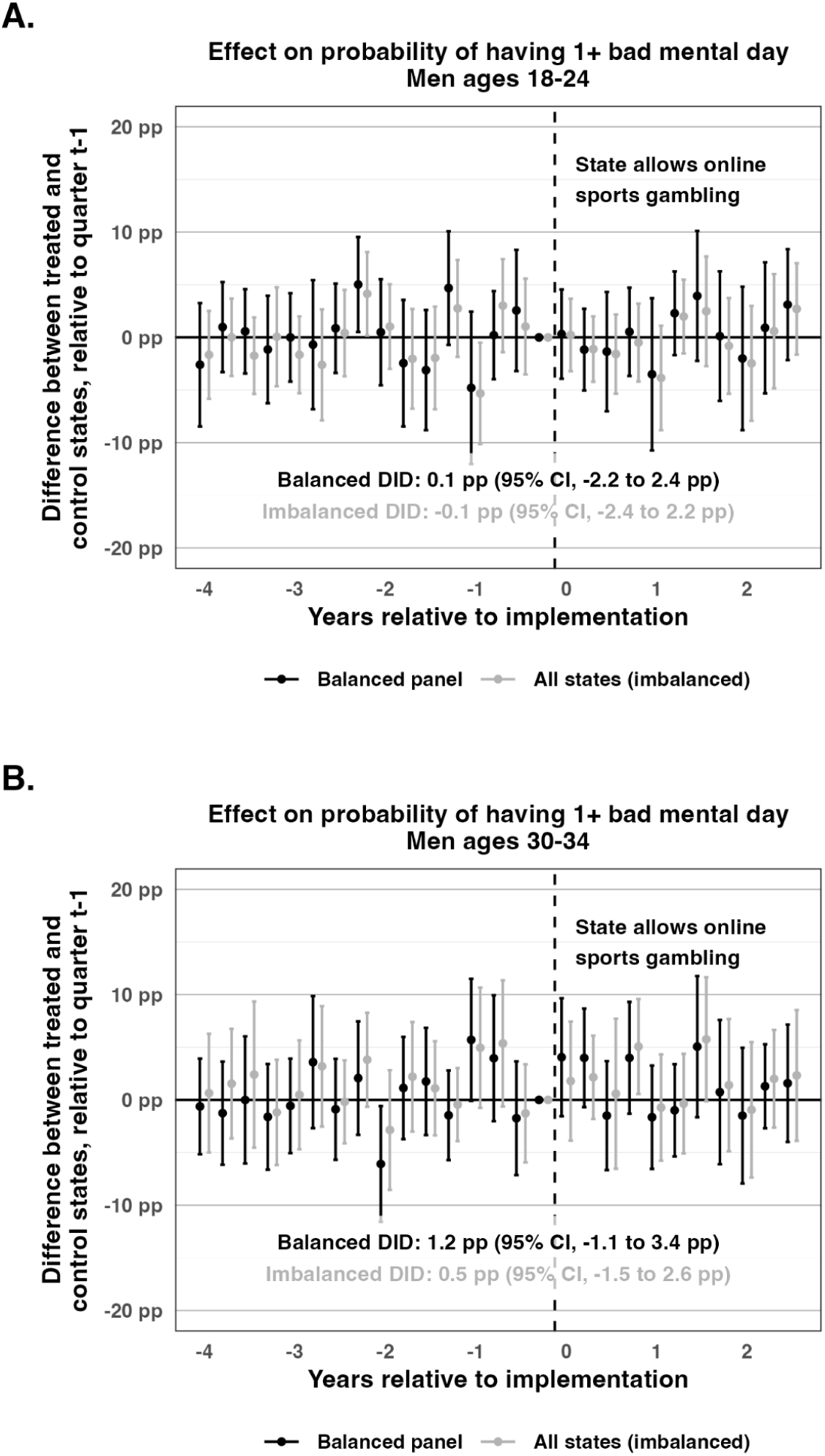
Replication of Couture et al (2024). **Notes:** We replicated the preferred outcome and age groups of Couture et al (2024), i.e. likelihood of having at least one bad mental health day among men ages 18–24 and 30–34. Models otherwise have the same specifications as our main models. We did not find clear evidence that online sports gambling affected the mental health of either group.

**eFigure 5.**
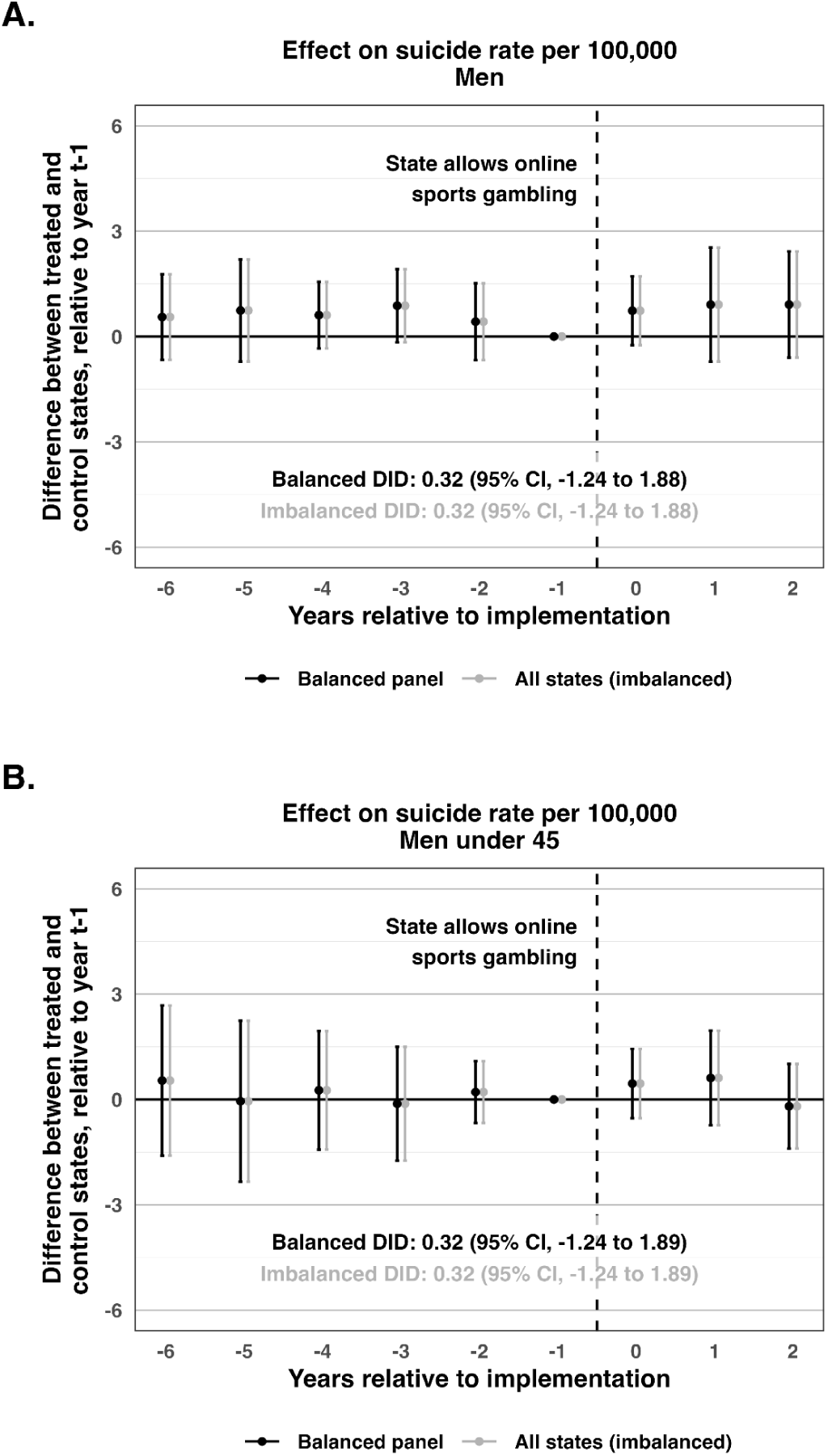
Event Studies for the Effect of Legalizing Online Sports Gambling on Suicide Mortality Among Men and Men Younger Than 45 Years. **Notes**: Based on CDC WONDER mortality records from 2012–2024. Both panels present stacked difference-in-differences event study estimates comparing treated and control states relative to the year before legalization of online sports gambling. **Panel A** shows suicide mortality per 100,000 population among men; **Panel B** shows suicide mortality among men younger than 45 years. Models included fixed effects for state-by-stack and year-by-stack. Estimates were weighted by population in each state-year. Standard errors were clustered by state, with cluster-robust 95% confidence intervals provided.

**eFigure 6.**
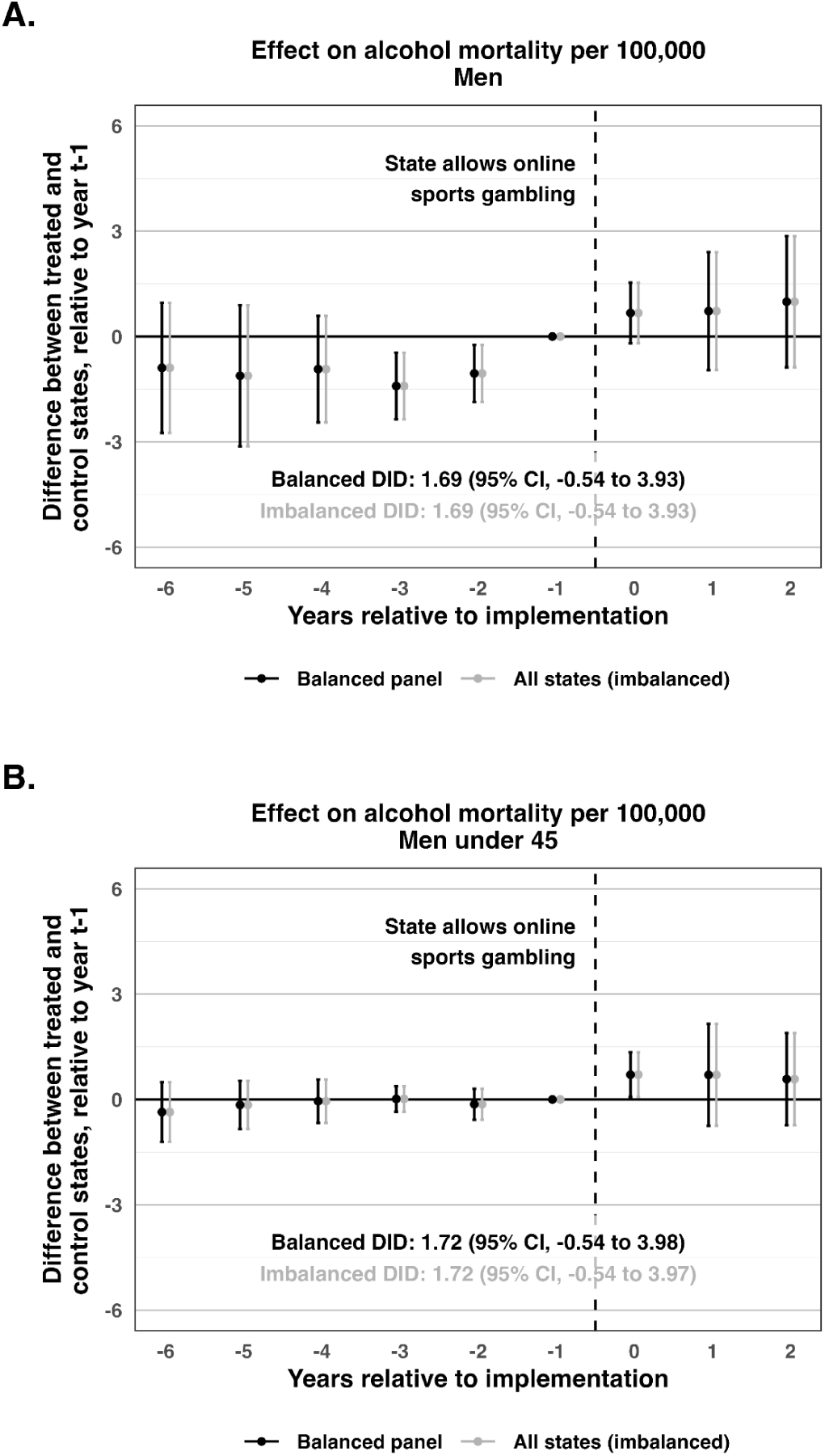
Event Studies for the Effect of Legalizing Online Sports Gambling on Alcohol-Induced Mortality Among Men and Men Younger Than 45 Years. **Notes**: Based on CDC WONDER mortality records from 2012–2024. Both panels present stacked difference-in-differences event study estimates comparing treated and control states relative to the year before legalization of online sports gambling. **Panel A** shows alcohol-induced mortality per 100,000 population among men; **Panel B** shows alcohol-induced mortality per 100,000 among men younger than 45 years. Models included fixed effects for state-by-stack and year-by-stack. Estimates were weighted by population in each state-year. 95% confidence intervals clustered at the state level are provided.

## eMethods: Stacked Difference-in-Differences Design

We used a stacked difference-in-differences design to estimate the causal effect of legalizing online sports gambling on population mental health (**eFigure 7**). This approach addresses potential biases in pooled difference-in-differences models when states adopt policies at different (or staggered) times and when treatment effects vary over time.

**eFigure 7.**
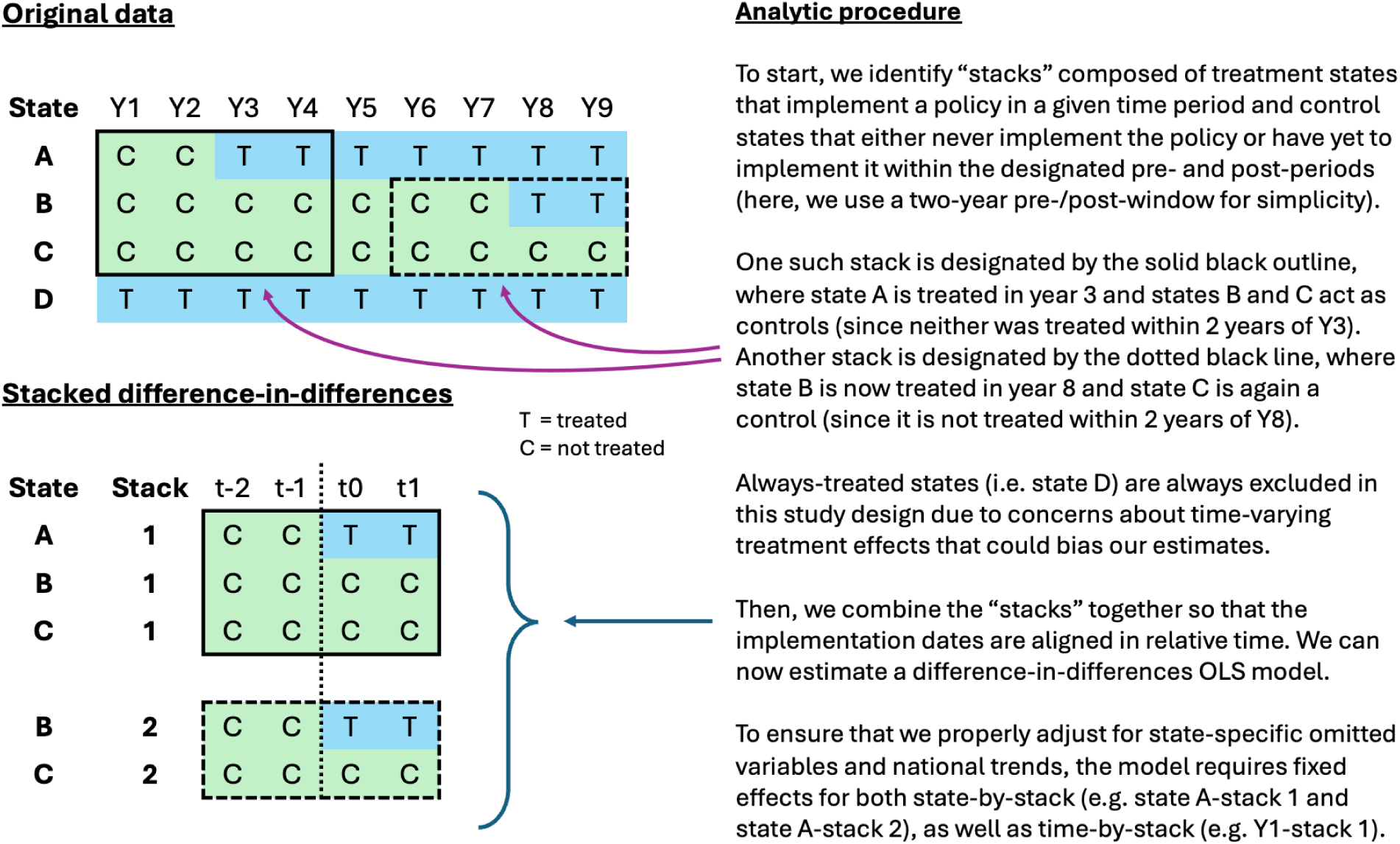
Schematic for a Sample Stacked Difference-in-Differences Design.

Rather than pooling all states into a single comparison, the stacked design allows us to isolate “stacks” of state(s) that implemented the policy at a given time and a counterfactual set of control states for that “stack.” We then align the timing within each stack relative to when the treatment state(s) legalized gambling and combine all “stacks” into one model.

Since each stack represents its own mini-experiment, the resulting difference-in-differences model requires fixed effects for state-by-stack (not just state) to eliminate state-level omitted variables within a given stack, as well as quarter-by-stack (not just quarter) to eliminate national trends within a stack. We also included fixed effects for respondents’ birth year to account for generational differences in the experience and reporting of mental health.

This approach ensures that we always make appropriate comparisons between treatment and control states and that we avoid using already-treated states as controls, which can bias estimates when there are heterogeneous treatment effects by time or place.

In the schematic, the panel of states is fully “balanced” in that all states have the full number of pre- and post-period years. In reality, several states have recently legalized online sports gambling and do not have a full 2.5 years of post-period data. Using imbalanced panels can introduce bias into difference-in-differences models, especially in the interpretation of event studies. For this reason, our main estimates are restricted to a balanced panel of 22 treated units. We also present an imbalanced panel of all 30 treated units in case states that recently legalized online gambling have experienced especially great mental health impacts. In practice, the point estimates of the two are virtually identical.

## Footnotes

^1^We base our definition of “young men” on birth years, rather than ages, to ensure that we follow the same cohorts of men over time. Conditioning on age would mean that each cross-sectional wave of the BRFSS examines a different and continuously shifting population. As such, it would be unclear whether any observed effects are attributable to gambling policy or differential demographic trends between treated and control states. In practice, however, the results were similar when specified either way.

